# Surgical Performance and Patient Outcomes in Robot-Assisted Esophagectomy: A Crowd-Sourced Video-Based Assessment

**DOI:** 10.64898/2026.09.25.26364010

**Authors:** Felix von Bechtolsheim, Nele Kolb, Sebastian Bodenstedt, Stefanie Speidel, Felix Merboth, Jasmin Hasanovic, Leonore Weikert, Olga Radulova-Mauersberger, Nathalie Buck, Felicitas La Roseé, Grit Krause-Jüttler, Jürgen Weitz, Marius Distler, Florian Oehme

## Abstract

**Background:** Robust evidence linking individual surgeon technical performance to patient outcomes in complex minimally invasive surgery remains scarce. Crowd-sourced video assessment offers a scalable alternative to expert review, but its clinical relevance in complex oncologic procedures such as robot-assisted minimally invasive esophagectomy (RAMIE) is unestablished.

**Methods:** In this exploratory study, video recordings of 60 consecutive RAMIE procedures performed by 7 surgeons (2018–2023) were segmented into five representative surgical steps and rated by 93 blinded crowd-raters per segment using a modified GEARS instrument. Tertiles were used to compare top-performing operations and steps against the remaining two terciles for overall and step-specific performance, and associations with perioperative outcomes were analyzed.

**Results:** Top-tercile overall performance was associated with significantly shorter operative time (abdominal, thoracic, and total) and less intraoperative blood loss (all p<0.05), without differences in postoperative outcomes. In exploratory step-specific analyses, top-tertile performance during infracarinal lymphadenectomy was associated with significantly fewer surgical complications (30.0% vs 60.0%; p=0.028), fewer severe surgical complications (25.0% vs 52.5%; p=0.043), and higher lymph node yield (22.8 vs 18.9; p=0.037), while the purse-string suture step showed the broadest operative-time benefit. These step-specific findings are hypothesis-generating and were not adjusted for multiple testing.

**Conclusion:** Crowd-sourced video assessment enables scalable, step-specific evaluation of surgical performance in RAMIE associated with intraoperative blood loss and operative time. Exploratory analyses suggest that the relationship between technical performance and clinical outcomes might be step-specific, with infracarinal lymphadenectomy emerging as risky step linked to postoperative morbidity. These findings support further investigation of individualized performance assessment for quality improvement and risk stratification.

## INTRODUCTION

Patient outcomes after complex surgery are influenced by a wide range of pre-, peri-, and postoperative factors. In contrast, the impact of the individual surgeon on the perioperative course remains far less well understood. Although surgical performance is widely presumed to be a major determinant of patient outcomes, the empirical evidence supporting this association remains scarce and heterogenous.^1,2^ This knowledge gap is particularly relevant in complex minimally invasive oncologic surgery, where a broad range of technical skills must be executed at a consistently high level, with their effective application being reflected in the surgeon’s overall performance and ultimately determining satisfactory outcomes.

Robot-assisted minimally invasive esophagectomy (RAMIE) exemplifies this challenge combining complex oncologic resection with demanding minimally invasive surgery. Variation in technical execution may plausibly translate into meaningful differences in postoperative morbidity, recovery, and oncologic quality.

While direct evidence for the impact of surgical performance in RAMIE is lacking, it can be assumed based on data showing that greater institutional experience and expertise with robot-assisted esophagectomy is associated with significant differences in postoperative morbidity and mortality.^3–5^ However, hospital-level outcomes reflect an entire care system and offer little insight into the individual surgeon’s contribution. Direct assessment of surgeon-level technical performance is therefore needed to better understand how intraoperative performance may influence outcomes.

Video-based assessment offers a promising approach to quantify and qualify surgical performance, particularly in robotic surgery, where high-quality operative recordings are routinely available. Expert review of intraoperative video can provide detailed evaluation of surgical performance, but this method is resource intensive and difficult to scale. Crowdsourced assessment has emerged as a potential alternative, using aggregated ratings from nonexpert reviewers to generate structured evaluations of operative skill.^6–8^ In other procedural settings, this approach has shown encouraging results, but its applicability to highly complex general surgery remains underused.^6,7^

Therefore, in this study, we aimed to utilize crowd-assessment to directly investigate the relationship between an individual surgeon’s performance and objective patient outcomes after RAMIE.

## MATERIAL AND METHODS

This exploratory, single-center trial combining retrospectively collected patient data with a video-based evaluation of surgical performance was approved by the local ethics committee of the TU Dresden (BO-EK-243052023) and registered at the German Clinical Trials Register (DRKS00040906).

### Study cohort and design

All consecutive Patients who underwent a robot-assisted minimally invasive esophagectomy (RAMIE) procedure at our institution between 2018 and 2023 were screened for the inclusion (Figure 1). Patients and their corresponding surgical recordings were eligible for inclusion if the procedure was completed as a standardized RAMIE without conversion to open or conventional laparoscopic surgery, and a complete video recording of the procedure was available

**Figure 1:**
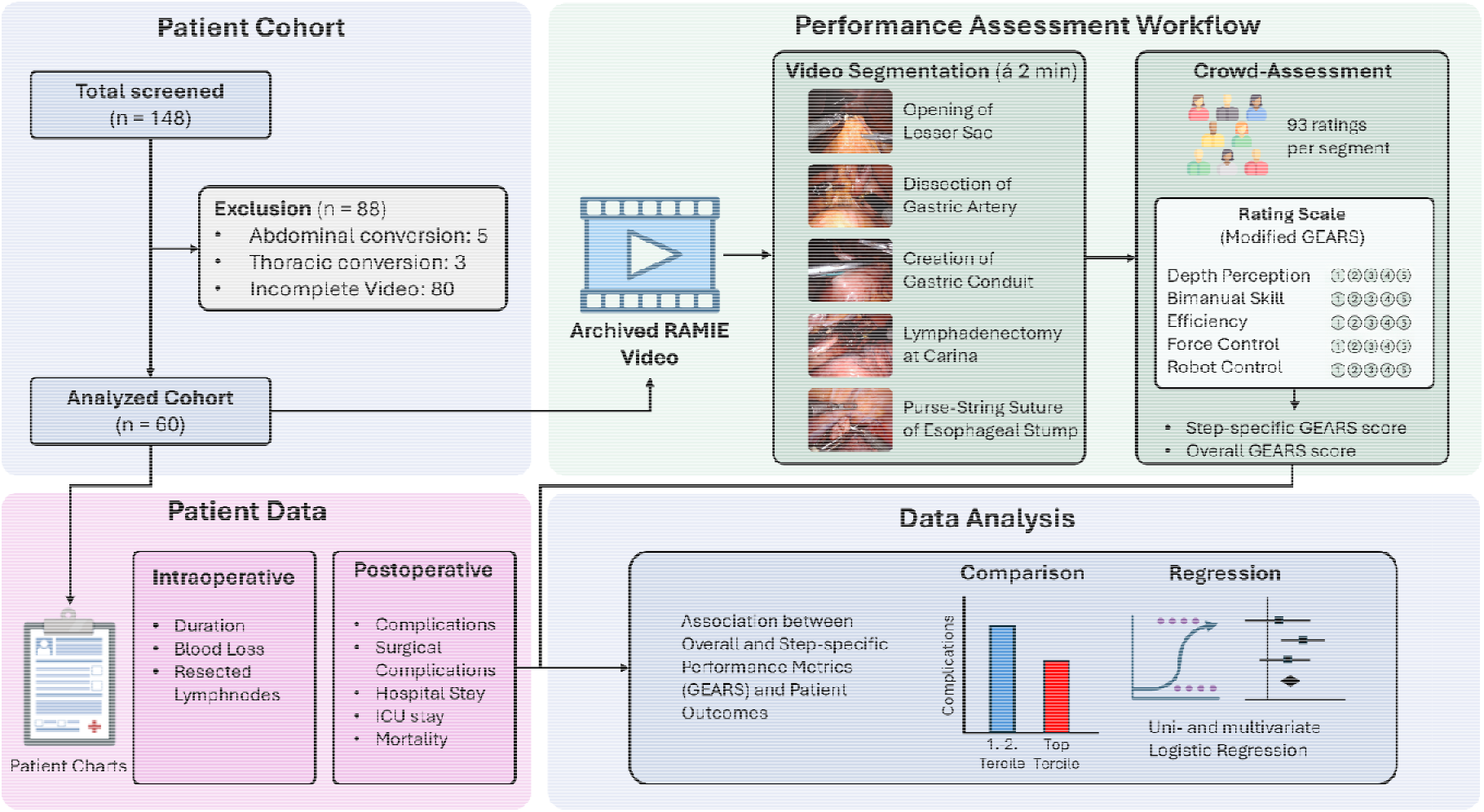
Trial scheme

#### Video-based performance assessment

The video material was compiled and segmented into five relevant surgical steps, which were selected by surgical experts as representative of their importance to the operation, as well as to cover a broad range of surgical activities. The five steps comprised:

- opening of the lesser sac
- dissection of left gastric artery
- creation of gastric conduit
- lymphadenectomy at the carina
- purse-string suture of the esophageal stump.

Each video segment was limited to exactly two minutes and was not further edited, in order to reduce the possible influence of the duration of a surgical procedure on the rating.

The video segments were anonymized and presented online to a pool of 93 randomly selected crowd-raters per segment (Clickworker GmbH, Essen, Germany) — laypersons naïve to surgery, reliability-checked and blinded to clinical data. Ratings used a modified GEARS instrument (five items: depth perception, bimanual skill, efficiency, force control, robot control) ^9^ on a 1–5 Likert scale, yielding a total score of 5–25 per segment; the overall GEARS score was calculated as the mean of the five segment scores.

#### Outcome Measures

Patient data were extracted from medical records, including intraoperative parameters (e.g., blood loss, operative duration) and postoperative course (complications, length of stay, mortality). Complications were graded using the Clavien–Dindo Classification and Comprehensive Complication Index (CCI)^10, 11^ with severe complications defined as CDC ≥ III and surgery-related complications (e.g., anastomotic leak, surgical site infection, pneumothorax) recorded separately from non-surgical complications.

The primary endpoint was the occurrence of postoperative surgical complications, compared between the top-performing tertile and the remaining two tertiles of overall and step-specific surgical performance. Secondary endpoints comprised the overall and severe complication rate, the comprehensive complication index, and quality indicators including blood loss, length of hospital stay, ICU stay, mortality, operative duration, and lymph node yield.

#### RAMIE procedure

All procedures were performed using a DaVinci Xi robotic system (Intuitive, Sunnyvale, CA, USA) with a standardized five-port configuration in the abdominal and thoracic phase. The abdominal phase started with opening of the lesser sac and gastric mobilization along the greater curvature, preserving the gastroepiploic arcade, followed by lymphadenectomy along standard lymph node stations and central division of the left gastric vessels. After mobilizing the distal esophagus through the hiatus, a gastric conduit was created along the lesser curvature using linear staplers.

Patients were then repositioned left lateral for the thoracic phase. The mediastinal pleura was then opened, the azygos vein divided, and the esophagus circumferentially mobilized from the diaphragmatic hiatus to the thoracic inlet combined with an *en bloc* lymphadenectomy of the periesophageal, subcarinal, and paratracheal nodal stations.

Following proximal esophageal transection via a linear stapler, the staple line was opened to place a continuous purse-string suture to secure the head-piece of a circular stapler. After exteriorizing the specimen through a mini-thoracotomy, an end-to-side esophagogastric anastomosis was constructed using the circular stapler. The remaining lesser curvature was divided, and the anastomotic staple line was reinforced with a running suture.

### Statistical analysis

SPSS version 29 (IBM Corp, Armonk NY, USA) was used for statistical analysis. Normality was tested via the Kolmogorov-Smirnov test. Variables are represented either as means with standard deviation (SD) for continuous variables or as frequencies (n) and percentage (%) for ordinal or categorial variables.

The mean total GEARS score (average across all five segments) was used to form a top-performing tertile, compared against the remaining two tertiles. For the step-specific analysis, tertiles were instead based on the GEARS score of that single step, so the same operation could fall into different tertiles depending on classification. Step-specific GEARS scores were related to perioperative outcomes recorded at the whole-operation level, rather than to step-specific outcomes.

For the statistical analysis of differences, a Student’s t test (for continuous variables) or Chi-squared test (for categorial or ordinal variables) was chosen. The threshold for the level of significance was defined as p < 0.05.

To account for multiplicity arising from testing the same outcome variables at multiple levels (overall cohort and five individual surgical segments), we distinguished between confirmatory and exploratory analyses. The primary endpoint — occurrence of surgical and severe surgical complications (Clavien–Dindo ≥ III) — was tested a priori only at the level of overall procedural performance. All step-specific analyses were pre-specified as exploratory, hypothesis-generating analyses intended to identify candidate high-risk steps for future confirmatory studies. Given the correlated, non-independent test structure (identical patients, overlapping outcomes) and limited sample size, no formal multiplicity correction was applied, as this would be overly conservative and risk obscuring clinically relevant signals. P-values for these step-specific comparisons are therefore reported descriptively and were not adjusted for multiplicity.

As an exploratory secondary analysis, a binary logistic regression model was additionally used to identify factors associated with the occurrence of surgical complications. Given the limited sample size, this analysis is reported in the Supplementary Material and should be interpreted as hypothesis-generating rather than confirmatory.

## RESULTS

### Patient characteristics

Between 2018 and 2023 148 patients received RAMIE. After screening, 8 patients were excluded due to conversion in the abdominal or thoracic phase whereas 80 patients had no or incomplete video recordings. In total 60 patients were eligible for inclusion. A total of 7 surgeons, all board-certified and experienced in robotic and esophageal surgery, performed all of the RAMIE procedures.

The patient cohort was predominantly male (90 %) and had a medium age of 62.7 (SD 8.8) years (Table 1). Most patients (76.7 %) were classified ASA 3, followed by ASA 2 (21.7 %). Smoking was the most frequent risk factor (43.3 %).

**Table 1:** Basic participant characteristics

| Basic Patient Characteristics |  |  |  |
| --- | --- | --- | --- |
| **Sex** | female | n (%) | 6 (10) |
|  | male |  | 54 (90) |
| **Age [years]** |  | mean (SD) | 63.5 (8.8) |
| **BMI [kg/m<sup>2</sup>]** |  | mean (SD) | 26.5 (4.7) |
| **ASA** | 1 | n (%) | 0 (0) |
|  | 2 |  | 13 (21.7) |
|  | 3 |  | 46 (76.7) |
|  | 4 |  | 1 (1.7) |
| **Smoking** |  | n (%) | 26 (43.3) |
| **Alcohol** |  | n (%) | 9 (15) |
| **Diabetes Mellitus** |  | n (%) | 10 (16.7) |
| **Histology** | Adeno-Carcinoma | n (%) | 44 (73.3) |
|  | Sqamous Carcinoma |  | 13 (21.7) |
|  | Undifferentiated Carcinoma |  | 1 (1.7) |
|  | Sarcoma |  | 1 (1.7) |
|  | Barrett's disease |  | 1 (1.7) |
| **UICC** | 0 | n (%) | 1 (1.7) |
|  | I |  | 14 (23.3) |
|  | IA |  | 6 (10) |
|  | IB |  | 6 (10) |
|  | II |  | 4 (6.7) |
|  | IIB |  | 2 (3.3) |
|  | III |  | 1 (1.7) |
|  | IIIA |  | 4 (6.7) |
|  | IIIB |  | 13 (21.7) |
|  | IVa |  | 6 (10) |
|  | IVB |  | 2 (3.3) |
| **Hospital stay [days]** |  | mean (SD) | 18.7 (14.5) |
| **ICU stay [days]** |  | mean (SD) | 4.4 (4.8) |
| **CCI (30 days)** |  | mean (SD) | 23.7 (22.8) |
| **Complications** |  | n (%) | 38 (63.3) |
| **Severe Complications** |  | n (%) | 31 (51.7) |
| **Surgical Complications** |  | n (%) | 30 (50) |
| **Severe Surgical Complications** |  | n (%) | 26 (43.3) |
| **Mortality (30 day)** |  | n (%) | 1 (1.7) |

Most patients had adenocarcinoma of the esophageal gastric junction or the esophagus (73.3 %) or squamous cell carcinoma (21.7 %). UICC stage I was the most common stage (43.4%), followed by UICC stage III (30.1 %).

### Overall Outcome Data

Overall, patients spent an average of 4.4 (SD 4.8) days on ICU and 18.7 (SD 14.5) days in the hospital. The mean comprehensive complication index was 23.7 (SD 22.8) and complications occurred in 38 (63.3 %) patients. A total of 31 (51.7 %) patients had severe complications (CDC ≥ III) whereas surgical complications and severe surgical complications (CDC ≥ III) occurred in 30 (50 %) and 26 (43.3 %) patients, respectively. One patient (1.7 %) died within 30 days. The most common complications were deep vein thrombosis (18.3 %) followed by delayed gastric emptying (16.7 %) and anastomotic leakage (11.7 %).

### Overall Performance Data

As stated in the statistical analysis section, the overall GEARS score (calculated as the mean GEARS score of all five segments) was split into a top performance tertile and two remaining tertiles, which were then compared. The top tertile achieved a mean overall GEARS score of 20.5 versus 19.6 for the lower tertiles (Supplementary Table 1). Segment-level scores (top tertile vs. lower tertiles) were: lesser sac opening, 21.1 vs. 19.6; left gastric artery dissection, 21.2 vs. 19.5; gastric conduit creation, 19.8 vs. 17.8; carinal lymphadenectomy, 21.4 vs. 19.7; and purse-string suturing, 21.8 vs. 19.8.

### Overall Group Performance and Perioperative Outcomes

#### Patient characteristics

No significant differences in patient characteristics were observed (Supplementary Table 2).

#### Operative outcomes

Top-tertile performers demonstrated significantly shorter operative times (Table 2). Mean abdominal phase duration was 171.5 (SD 31.7) min in the top tertile versus 203.9 (SD 54.2) min in the remaining patients (p = 0.017), and thoracic phase duration was similarly reduced (194.4 [SD 98.4] min vs. 246.2 [SD 62.8] min; p = 0.016). Total operative time was likewise significantly shorter (403.6 [SD 111.4] min vs. 488.4 [SD 107.6] min; p = 0.006). Mean intraoperative blood loss was also significantly lower in the top tertile (186.8 [SD 94.0] ml vs. 297.1 [SD 217.1] ml; p = 0.014).

**Table 2:** Comparison of Top tertile vs. 1. & 2. Tertile for the total GEARS score. Significant p-values are marked bold.

|  |  | Total GEARS |  | P-Value |
| --- | --- | --- | --- | --- |
|  |  | 1.-2. Tertile<br>n = 40 | Top Tertile<br>n = 20 |  |
| Operative Time Abdomen [min] | mean (SD) | 203.9 (54.2) | 171.5 (31.7) | **0.017** |
| Operative Time Thorax [min] | mean (SD) | 246.2 (62.8) | 194.4 (98.3) | **0.016** |
| Total Operative time [min] | mean (SD) | 488.4 (107.6) | 403.6 (111.4) | **0.006** |
| Blood loss [ml] | mean (SD) | 297.1 (217.1) | 186.8 (94) | **0.014** |
| Hospital stay [days] | mean (SD) | 20.4 (16.8) | 15.2 (8.6) | 0.194 |
| ICU stay [days] | mean (SD) | 4.4 (5.1) | 4.4 (4.2) | 0.985 |
| Lymph nodes resected [n] | mean (SD) | 19.6 (6.5) | 21.3 (10.6) | 0.517 |
| CCI (30 days) | mean (SD) | 21.7 (19.8) | 27.8 (28) | 0.319 |
| All Complications | n (%) | 26 (65) | 12 (60) | 0.705 |
| Severe Complications | n (%) | 20 (50) | 11 (55) | 0.584 |
| All Surgical Complications | n (%) | 21 (52.5) | 9 (45) | 0.584 |
| Severe Surgical Complications | n (%) | 17 (42.5) | 9 (45) | 0.854 |
| Mortality (30 day) | n (%) | 0 (0) | 1 (5) | 0.285 |

#### Postoperative outcomes

The primary endpoints, surgical complications and severe surgical complications, were comparable between groups. Length of hospital stay (15.2 [SD 8.6] days vs. 20.4 [SD 16.8] days; p = 0.194), ICU stay (4.4 days in both groups; p = 0.985), lymph node yield (21.3 [SD 10.6] vs. 19.6 [SD 6.5]; p = 0.517), and 30-day Comprehensive Complication Index (27.8 [SD 28.0] vs. 21.5 [SD 19.8]; p = 0.319) did not differ significantly between the top tertile and the remaining patients.

### Explorative Analysis of Step-Specific Surgical Performance and Perioperative Outcomes

#### Patient characteristics

Basic patient characteristics were comparable across step-specific tertile groups, with three exceptions (Supplementary Table 3): in the left gastric artery dissection step, the top tertile contained a higher proportion of smokers (32.5% vs. 65.0%; p = 0.017), and in the gastric conduit creation step, top-tertile patients were significantly older (66.7 vs. 61.9 years; p = 0.024). For the purse-string suture step, top-tertile patients had a significantly lower BMI (24.9 vs. 27.3 kg/m^2^; p = 0.03).

#### Operative outcomes

Step-specific top-tertile performance was associated with reduced intraoperative blood loss in the lesser sac opening step (187.5 [SD 114.6] ml vs. 300.0 [SD 213.2] ml; p = 0.016) (Table 3). For the gastric conduit creation step, thoracic phase duration was significantly shorter in the top tertile (203.9 min vs. 241.4 min; p = 0.043). In the purse-string suture step top-tertile performers achieved shorter times in the abdominal phase (174.1 [SD 32.7] min vs. 202.6 [SD 54.7] min; p = 0.036), thoracic phase (198.0 [SD 75.1] min vs. 244.4 [SD 78.0] min; p = 0.032), and total procedure (411.3 [SD 96.6] min vs. 484.6 [SD 117.0] min; p = 0.019). No significant differences in operative times or blood loss were found for the remaining steps.

**Table 3:** Step-specific comparison of Top Tertile vs. 1. & 2 for perioperative outcomes. P-values for these step-specific comparisons are therefore reported descriptively and were not adjusted for multiplicity, yet significant p-values are marked bold.

|  |  | Lesser sac opening |  |  | Dissection of gastric artery |  |  | Creation of Gastric Conduit |  |  | Lymphadenectomy at Carina |  |  | Suture of Esophageal Stump |  |  |
| --- | --- | --- | --- | --- | --- | --- | --- | --- | --- | --- | --- | --- | --- | --- | --- | --- |
|  |  | 1.-2.<br>Tertile | Top<br>Tertile | P-Value | 1.-2.<br>Tertile | Top<br>Tertile | P-Value | 1.-2.<br>Tertile | Top<br>Tertile | P-Value | 1.-2.<br>Tertile | Top<br>Tertile | P-Value | 1.-2.<br>Tertile | Top<br>Tertile | P-Value |
| **Operative Time** |  |  |  |  |  |  |  |  |  |  |  |  |  |  |  |  |
| **Abdomen [min]** | mean | 198.2 | 182.9 | 0.267 | 198.3 | 182.8 | 0.131 | 195.5 | 188.4 | 0.303 | 194.3 | 190.8 | 0.399 | 202.6 | 174.1 | **0.018** |
|  | (SD) | (41.7) | (63.6) |  | (54.7) | (38.3) |  | (47.1) | (56.5) |  | (53.7) | (43.1) |  | (54.7) | (32.7) |  |
| **Thorax [min]** | mean | 232.9 | 221 | 0.588 | 219.3 | 248.1 | 0.094 | 241.4 | 203.9 | **0.043** | 233.2 | 220.3 | 0.279 | 244.4 | 198 | **0.016** |
|  | (SD) | (74.1) | (91) |  | (72) | (91.9) |  | (68.8) | (94.5) |  | (84.5) | (69.8) |  | (78) | (75.1) |  |
| **Total [min]** | mean | 468.7 | 443 | 0.420 | 455.8 | 468.9 | 0.341 | 476.1 | 428.3 | 0.066 | 465.6 | 449.2 | 0.303 | 484.6 | 411.3 | **0.009** |
|  | (SD) | (103.2) | (137.5) |  | (115.6) | (116.9) |  | (105.3) | (129.8) |  | (117.8) | (111.9) |  | (117) | (96.6) |  |
| **Blood loss [ml]** | mean | 300 | 187.5 | **0.016** | 237.1 | 297.2 | 0.145 | 240 | 291.7 | 0.176 | 261.4 | 250 | 0.419 | 271.4 | 230.6 | 0.231 |
|  | (SD) | (213.2) | (114.6) |  | (184.8) | (196.6) |  | (201.4) | (162.9) |  | (176.6) | (216.9) |  | (199.7) | (169) |  |
| **Hospital stay [days]** | mean | 18.5 | 19.1 | 0.874 | 19.5 | 17 | 0.270 | 18.3 | 19.4 | 0.401 | 20.4 | 15.3 | 0.104 | 17.6 | 20.8 | 0.216 |
|  | (SD) | (14.8) | (15) |  | (16.8) | (9.6) |  | (13.9) | (16.6) |  | (16.1) | (11.1) |  | (13.4) | (17.4) |  |
| **ICU stay [days]** | mean | 4.4 | 4.5 | 0.970 | 3.8 | 5.8 | 0.063 | 4.8 | 3.8 | 0.224 | 4.8 | 3.7 | 0.191 | 4.3 | 4.8 | 0.352 |
|  | (SD) | (4.8) | (4.8) |  | (4.2) | (5.6) |  | (5.4) | (3) |  | (5.3) | (3.3) |  | (5.1) | (4.1) |  |
| **Lymph nodes resected [n]** | mean | 19.5 | 21.5 | 0.389 | 19.5 | 21.5 | 0.185 | 19.4 | 21.7 | 0.151 | 18.9 | 22.8 | **0.037** | 19.5 | 21.6 | 0.176 |
|  | (SD) | (7.3) | (9.5) |  | (8.5) | (7.2) |  | (7.4) | (9.3) |  | (7) | (9.6) |  | (6.7) | (10.3) |  |
| **CCI (30 days)** | mean | 24.3 | 22.6 | 0.795 | 21.6 | 27.9 | 0.158 | 22.6 | 25.9 | 0.298 | 26 | 19.1 | 0.134 | 21.1 | 29 | 0.105 |
|  | (SD) | (24.8) | (18.9) |  | (21.6) | (25) |  | (20.6) | (27.2) |  | (23.7) | (20.7) |  | (24.4) | (18.7) |  |
| **Complications** |  |  |  |  |  |  |  |  |  |  |  |  |  |  |  |  |
| **All** | n | 25 | 13 | 0.850 | 25 | 13 | 0.850 | 26 | 12 | 0.705 | 27 | 11 | 0.344 | 22 | 16 | 0.58 |
|  | (%) | (62.5) | (65) |  | (62.5) | (65) |  | (65) | (60) |  | (67.5) | (55) |  | (55) | (80) |  |
| **Severe** | n | 20 | 11 | 0.584 | 20 | 11 | 0.584 | 20 | 11 | 0.584 | 23 | 8 | 0.273 | 18 | 13 | 0.273 |
|  | (%) | (50) | (55) |  | (50) | (55) |  | (50) | (55) |  | (57.5) | (40) |  | (45) | (65) |  |
| **All Surgical** | n | 20 | 10 | 1 | 19 | 11 | 0.584 | 21 | 9 | 0.584 | 24 | 6 | **0.028** | 17 | 13 | 0.1 |
|  | (%) | (50) | (50) |  | (47.5) | (55) |  | (52.5) | (45) |  | (60) | (30) |  | (42.5) | (65) |  |
| **Severe Surgical** | n | 16 | 10 | 0.461 | 17 | 9 | 0.854 | 17 | 9 | 0.854 | 21 | 5 | **0.043** | 15 | 11 | 0.197 |
|  | (%) | (40) | (50) |  | (42.5) | (45) |  | (42.5) | (45) |  | (52.5) | (25) |  | (37.5) | (55) |  |
| **Mortality (30 day)** | n | 1 | 0 | 0.596 | 1 | 0 | 0.285 | 0 | 1 | 0.285 | 1 | 0 | 0.596 | 1 | 0 | 0.596 |
|  | (%) | (2.5) | (0) |  | (2.5) | (0) |  | (0) | (5) |  | (2.5) | (0) |  | (2.5) | (0) |  |

#### Postoperative outcomes

Top-tertile performance in the carinal lymphadenectomy step was associated with significantly fewer surgical complications (30.0% vs. 60.0%; p = 0.028), fewer severe surgical complications (25.0% vs. 52.5%; p = 0.043) (Figure 2), and a higher lymph node yield (22.8 vs. 18.9; p = 0.037). For all other steps, postoperative length of stay, ICU stay, lymph node yield, and CCI did not differ significantly between groups.

**Figure 2:**
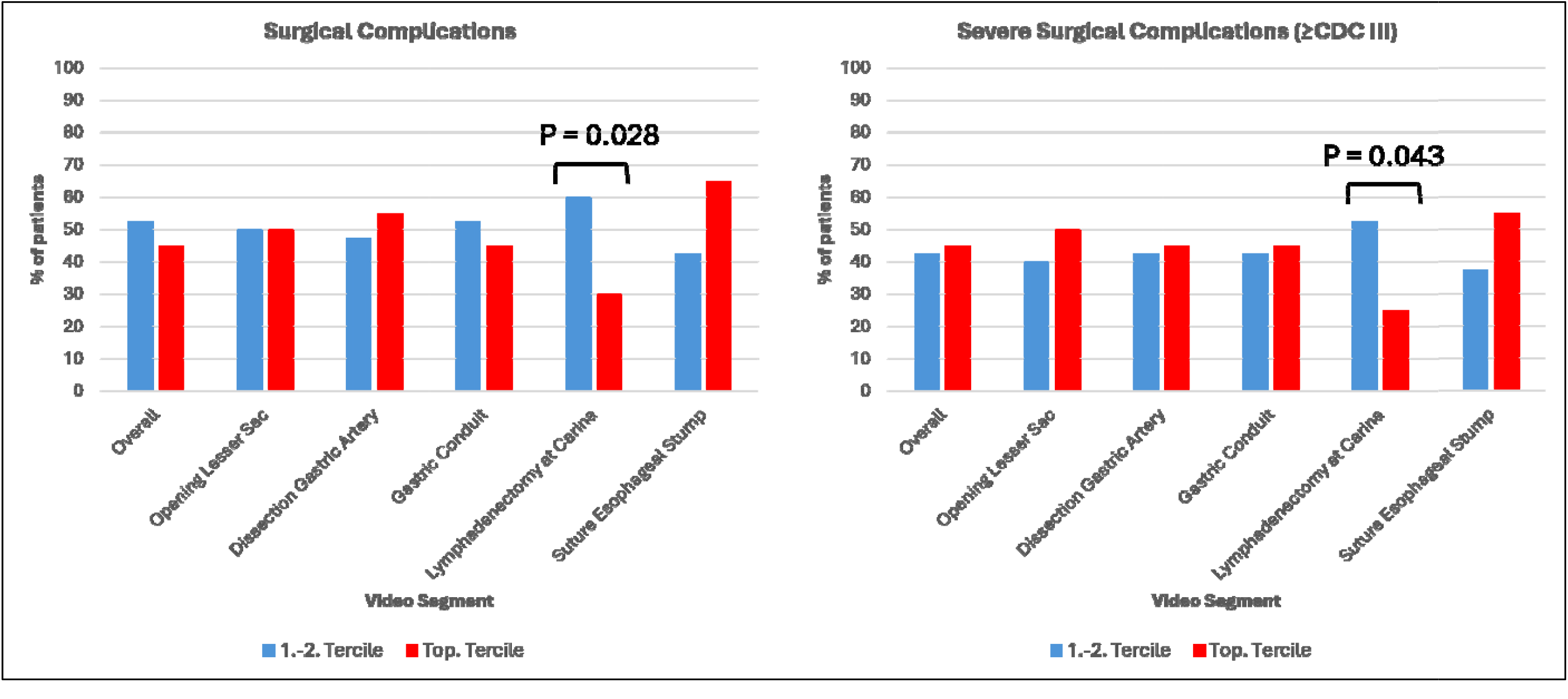
Comparison of surgical complications and severe surgical complications. P-values for these step-specific comparisons are therefore reported descriptively and were not adjusted for multiplicity.

### Regression analysis

In the uni- and multivariate analysis of a supplementary binary regression analysis only the GEARS score during the infracarinal lymphadenectomy (OR: 0.286; 95 % CI: 0.080 – 0.905; p = 0.034) and diabetes mellitus (OR: 0.075; 95 % CI: 0.009 – 0.669; p = 0.020) were significantly associated with a reduced risk of overall surgical complications (Supplementary Table 4).

## DISCUSSION

This study, including 60 patients undergoing RAMIE performed by 7 surgeons, demonstrates that crowd-sourced video assessment can granularly evaluate surgeon performance in highly complex oncological procedures like RAMIE, revealing that better intraoperative performance was associated with reduced operative time, lower blood loss, and – in exploratory step-specific analyses, during high-risk steps - a potential decrease of surgical complications.

In particular, better overall surgical performance was associated with shorter operating times and lower blood loss. In laparoscopic rectal resections and gastric bypasses, better surgical performance also led to a significantly shorter operative time and, in case of the rectal resections, also reduced blood loss.^12,13^ For robotic sleeve gastrectomies, no such association between surgical performance, indicated by GEARS, and operative time could been observed.^14^ This discrepancy suggests that the impact of surgical performance on operative duration is procedure-dependent, and might be more pronounced in complex, long-duration operations like RAMIE.

A hypothesis-generating observation is that the association between surgical performance and patient outcomes appears to vary by operative step – though, given the number of step-specific comparisons performed, these patterns should be regarded as exploratory rather than confirmed. Superior technical execution during the purse-string suture primarily optimized efficiency, shortening operative times, whereas higher GEARS scores during the infracarinal lymphadenectomy correlated with greater lymph node yield and reduced surgery-specific morbidity, roughly halving overall and severe complication rates in the top tertile. Notably, top-tertile performance during the lesser sac opening was associated with significantly lower intraoperative blood loss, raising the question of whether step-specific GEARS scores may carry predictive value for relevant intraoperative events, a hypothesis that warrants confirmation in adequately powered studies.

This pattern is consistent with Chhabra et al., who found that performance during proximal stomach dissection in bariatric sleeve gastrectomies correlated with clinical outcomes—including hemorrhage, leak rates, weight loss, and GERD severity—while other steps showed no association.^15^ This supports the notion that discrete procedural steps contribute unequally to outcomes, with the relevance for technical performance unevenly distributed across an operation.

This step-specific effect was further explored in our supplementary regression analysis, in which a higher GEARS score during infracarinal lymphadenectomy showed an independent association with fewer surgical complications (>70% risk reduction) — a finding that should be interpreted alongside the exploratory, uncorrected nature of the underlying step-specific comparisons, broadly consistent with prior work linking higher GEARS scores to better postoperative continence after robot-assisted prostatectomy and lower scores to greater complication risk after laparoscopic gastrectomy.^16,17^

Despite the clinical relevance of our findings, only few similar studies exist for direct comparison, underscoring the historical lack of scalability associated with structured surgical performance assessment. While expert assessment is considered the traditional gold standard, it is costly, time-consuming and lacks scalability. The present trial addressed this bottleneck by employing crowd assessment and is - to our knowledge - the first study to evaluate surgical performance using crowd-raters in robot-assisted esophageal surgery and to explore its association with postoperative outcomes.

In both simulated and clinical settings, crowd-assessment has proven to be an effective method for providing valid assessment of surgical skills and performance, matching expert feedback in several aspects.^6,7,18–21^ One trial including thoraco-laparoscopic esophagectomies reported heterogeneous results, likely due to its small sample of procedures and raters. However, Ketel et al. found a strong correlation between expert evaluations of entire videos and short clips,^22^ supporting the notion that representative key steps can capture surgical performance without requiring assessment of the entire procedure.

Postoperative morbidity after esophageal surgery, historically ranging between 43 % and 76 % - our cohorts profile matched these benchmarks - is directly linked to worse outcomes and reduced survival^23– 27x^. Anastomotic leaks in particular carry a 30-day mortality of 9% and a reduced long-term survival, more pronounced in low-volume than in high-volume hospitals.^3,28^

However, shifting the focus from broad institutional volume metrics to high-leverage technical steps allows for a more nuanced understanding of surgical outcomes. While hospital volume underscores a system’s capacity for crisis management, our findings might help to shift complication research away from describing hospital-level ‘failure to save’ toward optimizing surgeon-level ‘performance to prevent.’

Moving forward, this scalable evaluation framework could help targeted surgical education and quality improvement initiatives. Furthermore, integrating these crowd-assessment metrics into clinical risk-stratification models may hold potential for individualized patient outcome prediction, pending prospective evaluation in larger cohorts.

### Strengths and Limitations

Several limitations of the present trial warrant consideration, most notably our retrospective trial design and the modest sample size of 60 patients, which restricts statistical power. The counterintuitive diabetes-as-protective factor finding in the supplementary regression analysis likely reflects a statistical artifact or selection bias given cohort size. However, it is important to note that foundational studies investigating objective surgical skill have demonstrated robust, highly reproducible results with similarly sized cohorts.^17^

Furthermore, because all operations were performed by seven highly experienced, board-certified surgeons within a single specialized center, our findings may not reflect the steeper technical learning curves seen in broader, non-specialized settings. Conversely, the single-center design also controlled for confounding effects inherent to varying perioperative ecosystems of different hospitals, isolating the impact of individual technical performance.

A further limitation concerns multiple testing. We addressed this by pre-specifying a single confirmatory primary endpoint, tested only at the overall performance level, while all step-specific comparison were treated as exploratory from the outset. We therefore interpret the step-specific results, including the regression analysis, as hypothesis-generating rather than confirmatory. Validation in larger, prospectively designed cohort with pre-specified step-level endpoints is warranted.

### Conclusion

This trial indicates a relationship between intraoperative surgical performance and clinical relevant metrics in robot-assisted esophagectomy. Better surgical performance – particularly in specific steps – may be linked to fewer and less severe surgical complications, better lymph node yield, minimized blood loss, and shorter operative times.

Additionally, crowd-raters provide a valid and scalable methodology for objective surgical performance assessment. This may enable a potential paradigm shift from unspecific, hospital-level “failure to save” metrics to individual, surgeon-centric “performance to prevention,” a direction warranting further study towards personalized risk stratification and outcome prediction in complex minimally invasive oncology.

## Data Availability

The data supporting the findings of this study are not publicly available due to the sensitive nature of the research and privacy/ethical restrictions

