## Supplementary Material for "Surgical Performance and Patient Outcomes in Robot-Assisted Esophagectomy: A Crowd-Sourced Video-Based Assessment"

|  |  | 1.-2. Tertile | Top Tertile | P-Value |
| --- | --- | --- | --- | --- |
| **GEARS** | Overall | 19.6 | 20.5 | **<0.001** |
|  | Segment 1 | 19.6 | 21.1 | **<0.001** |
|  | Segment 2 | 19.5 | 21.2 | **<0.001** |
|  | Segment 3 | 17.8 | 19.8 | **<0.001** |
|  | Segment 4 | 19.7 | 21.4 | **<0.001** |
|  | Segment 5 | 19.8 | 21.8 | **<0.001** |

Supplementary Table 1: Comparison of GEARS scores overall and for each segment individually. Significant p-values are marked bold.

|  |  |  | **Total GEARS** | | |
| --- | --- | --- | --- | --- | --- |
|  |  |  | 1.-2. Tercile | Top Tercile |  |
|  |  |  | n = 40 | n = 20 | P-Value |
| **Sex** | female | n (%) | 5 (12.5) | 1 (5) | 0.361 |
|  | male |  | 35 (87.5) | 19 (95) |  |
| **Age [years]** |  | mean (SD) | 63.2 (8.7) | 64.1 (9.3) | 0.714 |
| **BMI [kg/m^2^]** |  | mean (SD) | 26.9 (4.9) | 25.6 (4.2) | 0.284 |
| **ASA** | 1 | n (%) | 0 (0) | 0 (0) |  |
|  | 2 |  | 6 (15) | 7 (35) | 0.174 |
|  | 3 |  | 33 (82.5) | 13 (65) |  |
|  | 4 |  | 1 (2.5) | 0 (0) |  |
| **Smoking** |  | n (%) | 17 (42.5) | 9 (45) | 0.854 |
| **Alcohol** |  | n (%) | 6 (15) | 3 (15) | 1 |
| **Diabetes Mellitus** |  | n (%) | 7 (17.5) | 3 (15) | 0.806 |
| **Histology** | Adeno-Carcinoma | n (%) | 28 (70) | 16 (80) | 0.111 |
|  | Sqamous Carcinoma | n (%) | 11 (27.5) | 2 (10) |  |
|  | Undifferentiated Carcinoma | n (%) | 0 (0) | 1 (5) |  |
|  | Sarcoma | n (%) | 0 (0) | 1 (5) |  |
|  | Barrett's disease | n (%) | 1 (2.5) | 0 (0) |  |
| **UICC** | 0 | n (%) | 1 (2.6) | 0 (0) | 0.488 |
|  | I |  | 8 (20.5) | 6 (30) |  |
|  | IA |  | 5 (12.8) | 1 (5) |  |
|  | IB |  | 5 (12.8) | 1 (5) |  |
|  | II |  | 2 (5.1) | 2 (10) |  |
|  | IIB |  | 2 (5.1) | 0 (0) |  |
|  | III |  | 1 (2.6) | 0 (0) |  |
|  | IIIA |  | 4 (10.3) | 0 (0) |  |
|  | IIIB |  | 8 (20.5) | 5 (25) |  |
|  | IVa |  | 2 (5.1) | 4 (20) |  |
|  | IVB |  | 1 (2.6) | 1 (5) |  |
| **Neoadjuvant Therapy** |  | n (%) | 32 (80) | 18 (90) | 0.327 |

Supplementary Table 2: Comparison of Top tertile vs. 1. & 2. Tertile for the basic patient characteristics.

|  |  |  | **Lesser sac opening** | | | **Dissection of gastric artery** | | | **Creation of Gastric Conduit** | | | **Lymphadenectomy at Carina** | | | **Suture of Esophageal Stump** | | |
| --- | --- | --- | --- | --- | --- | --- | --- | --- | --- | --- | --- | --- | --- | --- | --- | --- | --- |
|  |  |  | 1.-2. Tercile | Top Tercile |  | 1.-2. Tercile | Top Tercile |  | 1.-2. Tercile | Top Tercile |  | 1.-2. Tercile | Top Tercile |  | 1.-2. Tercile | Top Tercile |  |
|  |  |  | n = 39 | n = 20 | P-Value | n = 40 | n = 20 | P-Value | n = 40 | n = 20 | P-Value | n = 40 | n = 20 | P-Value | n = 40 | n = 20 | P-Value |
| **Sex** | female | n (%) | 5 (12.5) | 1 (5) | 0.361 | 5 (12.5) | 1 (5) | 0.361 | 5 (12.5) | 1 (5) | 0.361 | 4 (10) | 2 (10) | 1 | 2 (5) | 4 (20) | 0.068 |
|  | male |  | 35 (87.5) | 19 (95) |  | 35 (87.5) | 19 (95) |  | 35 (87.5) | 19 (95) |  | 36 (90) | 18 (90) |  | 38 (95) | 16 (80) |  |
| **Age [years]** | | mean (SD) | 63.1 (8.2) | 64.3 (10.2) | 0.616 | 63.7 (8.9) | 63.2 (9) | 0.834 | 61.9 (8.9) | 66.7 (8.1) | **0.024** | 63.6 (8.6) | 63.4 (9.6) | 0.946 | 63.9 (9.1) | 62.9 (8.5) | 0.344 |
| **BMI [kg/m^2^]** | | mean (SD) | 26.4 (4.9) | 26.7 (4.3) | 0.846 | 27.1 (4.7) | 25.3 (4.4) | 0.15 | 26.3 (4.9) | 26.8 (4.3) | 0.37 | 26.5 (4.6) | 26.5 (4.8) | 0.972 | 27.3 (4.6) | 24.9 (4.4) | **0.03** |
| **ASA** | 1 | n (%) | 0 (0) | 0 (0) | 0.125 | 0 (0) | 0 (0) | 0.748 | 0 (0) | 0 (0) | 0.5 | 0 (0) | 0 (0) | 0.717 | 0 (0) | 0 (0) | 0.062 |
|  | 2 |  | 11 (27.5) | 2 (10) |  | 9 (22.5) | 4 (20) |  | 10 (25) | 3 (15) |  | 8 (20) | 5 (25) |  | 6 (15) | 7 (35) |  |
|  | 3 |  | 29 (72.5) | 17 (85) |  | 30 (75) | 16 (80) |  | 29 (72.5) | 17 (85) |  | 31 (77.5) | 15 (75) |  | 34 (85) | 12 (60) |  |
|  | 4 |  | 0 (0) | 1 (5) |  | 1 (2.5) | 0 (0) |  | 1 (2.5) | 0 (0) |  | 1 (2.5) | 0 (0) |  | 0 (0) | 1 (5) |  |
| **Smoking** | | n (%) | 19 (47.5) | 7 (35) | 0.357 | 13 (32.5) | 13 (65) | **0.017** | 19 (47.5) | 7 (35) | 0.357 | 15 (37.5) | 11 (55) | 0.197 | 17 (42.5) | 9 (45) | 0.854 |
| **Alcohol** | | n (%) | 7 (17.5) | 2 (10) | 0.443 | 6 (15) | 3 (15) | 1 | 7 (17.5) | 2 (10) | 0.443 | 4 (10) | 5 (25) | 0.125 | 7 (17.5) | 2 (10) | 0.443 |
| **Diabetes Mellitus** | | n (%) | 7 (17.5) | 3 (15) | 0.806 | 7 (17.5) | 3 (15) | 0.806 | 5 (12.5) | 5 (25) | 0.221 | 6 (15) | 4 (20) | 0.624 | 9 (22.5) | 1 (5) | 0.086 |
| **Histology** | Adeno-Carcinoma | n (%) | 28 (71.8) | 16 (80) | 0.111 | 31 (79.5) | 13 (65) | 0.295 | 28 (70) | 16 (84.2) | 0.202 | 26 (66.7) | 18 (90) | 0.264 | 32 (80) | 12 (63.2) | 0.238 |
|  | Sqamous Carcinoma |  | 11 (28.2) | 2 (10) |  | 7 (17.9) | 6 (30) |  | 11 (27.5) | 2 (10.5) |  | 11 (28.2) | 2 (10) |  | 6 (15) | 7 (36.8) |  |
|  | Undiff. Carcinoma |  | 0 (0) | 1 (5) |  | 1 (2.6) | 0 (0) |  | 0 (0) | 1 (5.3) |  | 1 (2.6) | 0 (0) |  | 1 (2.5) | 0 (0) |  |
|  | Sarcoma |  | 0 (0) | 1 (5) |  | 0 (0) | 1 (5) |  | 1 (2.5) | 0 (0) |  | 1 (2.6) | 0 (0) |  | 1 (2.5) | 0 (0) |  |
|  | Barrett's disease |  | 0 (0) | 0 (0) |  | 1 (2.6) | 0 (0) |  | 0 (0) | 1 (5.3) |  | 1 (2.6) | 0 (0) |  | 0 (0) | 1 (5.3) |  |
| **UICC** | 0 | n (%) | 1 (2.6) | 0 (0) | 0.199 | 1 (2.6) | 0 (0) | 0.860 | 1 (2.5) | 0 (0) | 0.480 | 0 (0) | 1 (5) | 0.821 | 1 (2.5) | 0 (0) | 0.444 |
|  | I |  | 7 (17.9) | 7 (35) |  | 9 (23.1) | 5 (25) |  | 9 (22.5) | 5 (26.3) |  | 11 (28.2) | 3 (15) |  | 7 (17.5) | 7 (36.8) |  |
|  | IA |  | 5 (12.8) | 1 (5) |  | 3 (7.7) | 3 (15) |  | 2 (5) | 4 (21.1) |  | 5 (12.8) | 1 (5) |  | 5 (12.5) | 1 (5.3) |  |
|  | IB |  | 5 (12.8) | 1 (5) |  | 4 (10.3) | 2 (10) |  | 5 (12.5) | 1 (5.3) |  | 4 (10.3) | 2 (10) |  | 3 (7.5) | 3 (15.8) |  |
|  | II |  | 4 (10.3) | 0 (0) |  | 2 (5.1) | 2 (10) |  | 2 (5) | 2 (10.5) |  | 2 (5.1) | 2 (10) |  | 3 (7.5) | 1 (5.3) |  |
|  | IIB |  | 2 (5.1) | 0 (0) |  | 1 (2.6) | 1 (5) |  | 2 (5) | 0 (0) |  | 1 (2.6) | 1 (5) |  | 2 (5) | 0 (0) |  |
|  | III |  | 0 (0) | 1 (5) |  | 1 (2.6) | 0 (0) |  | 1 (2.5) | 0 (0) |  | 1 (2.6) | 0 (0) |  | 1 (2.5) | 0 (0) |  |
|  | IIIA |  | 4 (10.3) | 0 (0) |  | 4 (10.3) | 0 (0) |  | 2 (5) | 2 (10.5) |  | 3 (7.7) | 1 (5) |  | 4 (10) | 0 (0) |  |
|  | IIIB |  | 8 (20.5) | 5 (25) |  | 8 (20.5) | 5 (25) |  | 9 (22.5) | 4 (21.1) |  | 7 (17.9) | 6 (30) |  | 10 (25) | 3 (15.8) |  |
|  | IVa |  | 2 (5.1) | 4 (20) |  | 4 (10.3) | 2 (10) |  | 5 (12.5) | 1 (5.3) |  | 4 (10.3) | 2 (10) |  | 3 (7.5) | 3 (15.8) |  |
|  | IVB |  | 1 (2.6) | 1 (5) |  | 2 (5.1) | 0 (0) |  | 2 (5) | 0 (0) |  | 1 (2.6) | 1 (5) |  | 1 (2.5) | 1 (5.3) |  |
| **Neoadjuvant Therapy** | | n (%) | 33 (82.5) | 17 (85) | 0.806 | 31 (77.5) | 19 (95) | 0.086 | 34 (85) | 16 (80) | 0.624 | 34 (85) | 16 (80) | 0.624 | 34 (85) | 16 (80) | 0.624 |

Supplementary Table 3: Table 3: Step-specific comparison of Top Tertile vs. 1. & 2. Tertile for basic patient characteristics. P-values for these step-specific comparisons are therefore reported descriptively and were not adjusted for multiplicity, yet significant p-values are marked bold.

| **Binary regression analysis** | | **Univariate** | | | **Multivariate** | | |
| --- | --- | --- | --- | --- | --- | --- | --- |
|  |  | **Odds Ratio** | **95% CI** | **P-value** | **Odds Ratio** | **95% CI** | **P-value** |
| Age | | 1.027 | 0.97 - 1.09 | 0.369 |  |  |  |
| Sex | | 0.172 | 0.02 - 1.58 | 0.119 |  |  |  |
| ASA | | 1.415 | 0.44 - 4.52 | 0.558 |  |  |  |
| Smoking | | 0.438 | 0.15 - 1.24 | 0.121 |  |  |  |
| Alcohol | | 0.235 | 0.04 - 1.24 | 0.088 |  |  |  |
| Diabetes Mellitus | | 0.080 | 0.01 - 0.68 | **0.021** | 0.075 | 0.01 - 0.67 | **0.020** |
| BMI | | 1.015 | 0.91 - 1.13 | 0.792 |  |  |  |
| UICC | | 0.870 | 0.74 - 1.03 | 0.098 |  |  |  |
| Neoadj. Therapy | | 0.804 | 0.38 - 1.7 | 0.569 |  |  |  |
| GEARS | Gastrocolic Ligament | 1.000 | 0.34 - 2.93 | 1.000 |  |  |  |
|  | LAD Celiac trunc | 1.351 | 0.46 - 3.97 | 0.584 |  |  |  |
|  | Gastric Sleeve | 0.740 | 0.25 - 2.17 | 0.584 |  |  |  |
|  | LAD infracarinal | 0.286 | 0.09 - 0.9 | **0.032** | 0.268 | (0.08 - 0.9) | **0.034** |
|  | Purse String | 2.513 | 0.83 - 7.64 | 0.105 |  |  |  |
|  | Total | 0.740 | 0.25 - 2.17 | 0.584 |  |  |  |

Supplementary Table 4: A binary logistic regression model (univariate and multivariate) was used to identify risk or protective factors for the occurrence of surgical complications. Variables reaching a significance level of p < 0.05 in univariate analysis, were included in the multivariate model. The threshold for statistical significance was defined as p < 0.05. Significant p-values are marked bold.
